# Beyond Anxiety and Grief: Mapping the Emotional Landscape of Parents Facing a Childhood Cancer Diagnosis

**DOI:** 10.1101/2023.05.24.23290421

**Authors:** Shanzeh Sheikh, Courtney E. Wimberly, Lisa Towry, Kyle M. Walsh

## Abstract

**Objective:** We sought to explore the variation in emotional responses and identify clusters of emotional patterns associated with sociodemographic, clinical, and familial factors.

**Methods:** A large-scale survey with questions on demographics, experiences, and emotions at the time of diagnosis was sent to childhood cancer caregivers and completed between August 2012 and April 2019. Dimensionality reduction and statistical tests for independence were used to investigate relationships between sociodemographic, clinical, and psychosocial factors and 32 representative emotions.

**Results:** Data from 3142 respondents were analyzed. Through principal components analysis and t-distributed stochastic neighbor embedding analysis, three clusters of emotional responses were identified, captured 44%, 20% and 36% of respondents, respectively. Hallmark emotions within each cluster were “anger and grief” (Cluster 1), “pessimism, relief, impatience, insecurity, discouragement, and calm” (Cluster 2), and “hope” (Cluster 3). Cluster membership was associated with differences in parental factors, such as educational attainment, family income, and biological parent status, as well as child-specific factors, including age at diagnosis and cancer type.

**Conclusions:** The study revealed substantial heterogeneity in emotional responses to a child’s cancer diagnosis than previously recognized, with differences linked to both caregiver and child-related factors. These findings underscore the importance of developing responsive and effective programs to improve targeted support for caregivers from the time of diagnosis throughout a family’s childhood cancer journey.

## INTRODUCTION

Each year, about 16,000 children and adolescents are diagnosed with cancer in the United States.^1^ While improved treatments now enable over 80% of U.S. childhood cancer patients to survive long-term, cancer remains the leading cause of death by disease for children in the U.S.^2^ A childhood cancer diagnosis can be devastating for the patient and the family. Parents may struggle to cope, experiencing a range of negative emotions including increased anxiety and anticipatory grief, and this distress may be compounded by the processes of mourning and psychological preparations for a child’s possible death.^3^ However, emotional responses to a child’s cancer diagnosis are not uniform across families, and even within families, mothers and fathers have be observed to experience a child’s diagnosis differently.^4^ The nature of heterogeneity in parental emotional response to a childhood cancer diagnosis has been inadequately characterized, as have the roles of intrinsic and extrinsic factors contributing to such differences (*e.g.*, sociodemographic factors, family dynamics, childhood cancer subtypes).

Parental well-being is an important factor in considering caregivers as individuals with their own physical and psychological needs. Parental well-being is critical to childcare provision, particularly when a family faces a devastating medical diagnosis. Prior research indicates that parents of children newly diagnosed with cancer report high levels of personal sacrifice, sadness, and worry for the future,^3^ and that insufficient positive coping is associated with heightened anticipatory grief in family caregivers of adult cancer patients.^5^ Fear of a negative prognosis, difficulties with familial relationships, and financial hardship may further exacerbate parental psychological distress at diagnosis and in the years following.^6,7,8,9^

Negative impacts of a childhood cancer diagnosis on the family unit are clear, but existing data are primarily drawn from small, single-institution studies of patients treated at major academic medical centers. Furthermore, such studies often fail to explore the full emotional spectrum beyond the classic triad of anxiety, depression, and anticipatory grief. Such studies may overlook the role of other important post-diagnosis emotions, both positive (*e.g.*, optimism, empowerment) and negative (*e.g.*, guilt, frustration). Studies in parents of adolescents and young adults with advanced cancer have described how parental emotions fluctuate and transform over the course of their journey, relating the emergence of control, acceptance, hope, and support.^10,11^ Additional research is necessary to understand the complex range of emotions elicited by a childhood cancer diagnosis and to identify patient and family-specific factors contributing to differences in parental well-being.

To better understand the emotional state of parents at the time of a child’s cancer diagnosis and to understand sources of heterogeneity in their emotional response, we partnered with Alex’s Lemonade Stand Foundation’s (ALSF) to collect and analyze survey data from more than 3100 families affected by a childhood cancer diagnosis. We explored variation in parental emotions following a child’s cancer diagnosis to broadly quantify similarities and differences across respondents. We also explored how parental emotions were associated with a variety of sociodemographic, clinical, and psychosocial factors. The results of this study may help identify factors associated with poorer emotional well-being, with important implications for creating responsive and effective programs to provide targeted support to parents at an early point during a family’s childhood cancer journey.

## METHODS

### Study Population

To explore associations between parent/caregiver emotional state following a child’s cancer diagnosis, we partnered with ALSF to conduct an ongoing series of longitudinal surveys of families affected by childhood cancer. From 2011 to 2022, the ALSF My Childhood Cancer (MCC): Survey Series explored families’ experiences and attitudes from diagnosis, throughout treatment and follow-up care, and after bereavement (when applicable).^12^ MCC was an English-language survey series publicly hosted on the ALSF website and advertised via Facebook, Twitter, and the ALSF childhood cancer listserv. MCC targeted parental respondents (including step-parents and adoptive parents) whose child was diagnosed with cancer before his or her 18th birthday. Participation in MCC was not limited by the child’s current age, only their age at diagnosis. 3150 families participated in the MCC survey series.

In this cross-sectional study, we examined responses to the ALSF MCC diagnosis survey completed between August 2012 and April 2019 (N = 3142 respondents). The number of families in which two parents independently completed the survey was too small to make within-family comparisons, so the analyses presented are limited to one survey response per family. For families recording responses from multiple parents, the survey that was more complete was retained. If both surveys were fully completed, then the first survey returned was included in analyses. This study was approved by the Duke University Institutional Review Board (Pro00100771).

### Survey instruments

Childhood cancer type and patient/parental demographics were collected during MCC: Survey Series registration. Due to the low number of participants who identified as belonging to a racial/ethnic group other than non-Hispanic white, respondent race/ethnicity was collapsed into an indicator for “non-Hispanic white” and “Other.” Birth order was collapsed into “only child” versus “siblings.” Household income was recorded in the following bins: <$50,000; $50,000–$99,999; $100,000+ and modeled as an ordinal variable, with those answering and “prefer not to say” excluded from analysis. Marital status was collapsed into “married/living with domestic partner” versus “divorced, widowed, separated, or never married”. Cancer type was analyzed as “CNS” (central nervous system), “hematologic” (including leukemias and lymphomas), or “other solid tumor”. For all variables, respondents answering “unsure” were excluded from analysis.

Within the attitudes, anxieties, and emotions portion of the MCC Diagnosis Survey, respondents were presented with a list of 32 emotions. From this list, participants were asked to select up to 10 emotions that best reflected their feelings at the time of their child’s cancer diagnosis. To reduce biases related to the order in which they appeared, emotions were dynamically randomized for each respondent. Following dimensionality reduction (described below), respondent membership within identified clusters of emotions were treated as the primary dependent variable in downstream analyses.

Independent variables were derived from survey responses. Parental variables included: respondent sex, race/ethnicity, whether they were a biological parent of the child, education level, marital status, time from child’s diagnosis to survey completion, parental age at child’s diagnosis, insurance status, household income, locale (urban, suburban, rural), and the number of other children in the household. Variables pertaining to the child included: sex, age at diagnosis (including a binary category for infantile-onset cancers), whether the child had siblings, and cancer type (hematologic, CNS, or other solid tumor). Missing data rates for variables were very low, ≤1% for all except locale, which had a missing data rate of 6%. If data were missing for a modeled covariate, the individual with missing data was excluded from the model.

### Statistical Analyses

For analysis, each of the 32 emotions was coded as a dichotomous 0/1 value that indicated whether it was selected (1) or not selected (0) by a given respondent as reflecting their feelings at the time of their child’s cancer diagnosis. Because of the large number of emotions assessed and their highly-correlated nature, dimensionality reduction techniques were performed on these variables. First, we explored the data using principal components analysis to visualize the data in 2-dimensional space and to quantify the percent of variance explained by individual principal components (PCs) using the R package prcomp. Next, we applied t-distributed stochastic neighbor embedding (t-SNE) analysis to the 32 emotion variables using the package Rtsne (version 0.15) and a perplexity ranging from 10-50, with respondent values plotted in three dimensions.^13^ Finally, using the coordinates of individual respondents in three-dimensional t-SNE space, K-means clustering was used to identify any clusters and to assign respondents to membership within specific clusters (as t-SNE performs clustering but not classification). The number of clusters was iterated from K=1 up to K=10, with the sum of squared distances of each point from its centroid plotted against the value of K. The optimal K value was selected using the elbow method, where both the sum of squared distances and the number of clusters is minimized. Clusters were visualized using scatterplot3d (version 0.3.41).^14^

Relationships between independent and dependent variables (emotion clusters) were assessed using Chi-square tests for independence or Kruskal-Wallis tests, depending on whether the independent data were categorical or ordinal, respectively. For analysis of child age at diagnosis, a Box-Cox transformation was applied (MASS version 7.3-54)^15^ and ANOVA test performed. For all statistical association tests, α = 0.05 was used to determine nominal statistical significance. R Statistical Software (version 4.1.1)^16^ was used for statistical analyses.

## RESULTS

### Study Population

Our study sought to explore associations between parent/caregiver emotional state following a child’s cancer diagnosis, as well as child-specific and parent/caregiver-specific factors. Between August 2012 and April 2019, a total of 3142 respondents from unique families completed the baseline diagnosis survey containing questions about emotions following a child’s cancer diagnosis. Respondents were majority female (94%), non-Hispanic white (89%), the biological parent of the child diagnosed with cancer (98%), and married or living with their domestic partner (87%) (**Table 1**). Slightly more than half of respondents lived in suburban areas (56%). Household incomes were broadly distributed, with 30% of respondents earning <$50,000 annually, 41% earning $50-99,000 annually, 24% earning >$100,000 annually, and the remaining 5% missing or preferring not to respond. Slightly more than half of the respondents’ children were male (55%), and the median age of the child at time of diagnosis was 4 years (IQR 2-9). The median time from the child’s diagnosis to survey completion was 3 years (IQR 1-7). 42% of respondents’ children were diagnosed with a hematologic cancer, 40% were diagnosed with a non-CNS solid tumor, and 18% were diagnosed with a central nervous system (CNS) tumor.

**Table 1.**
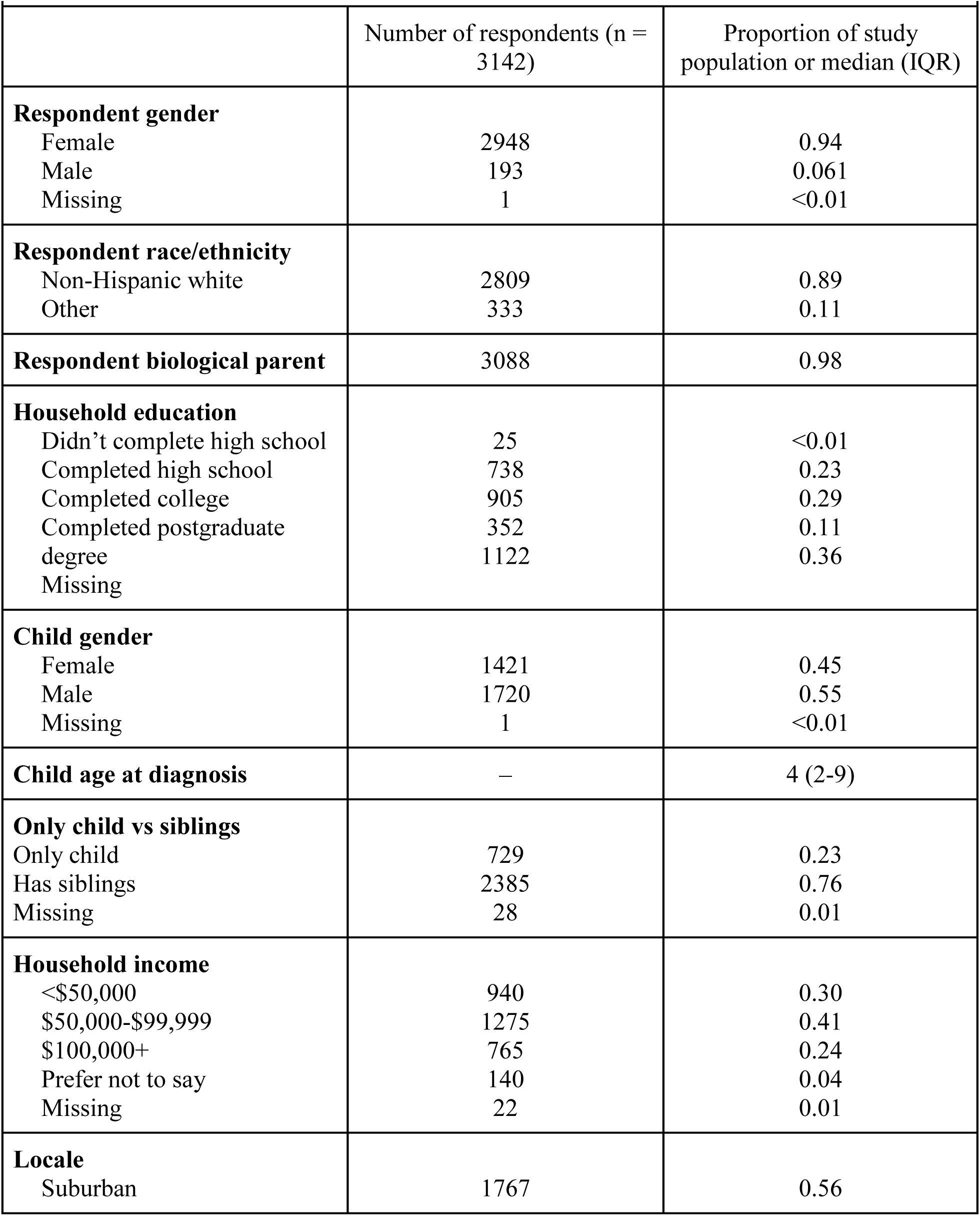
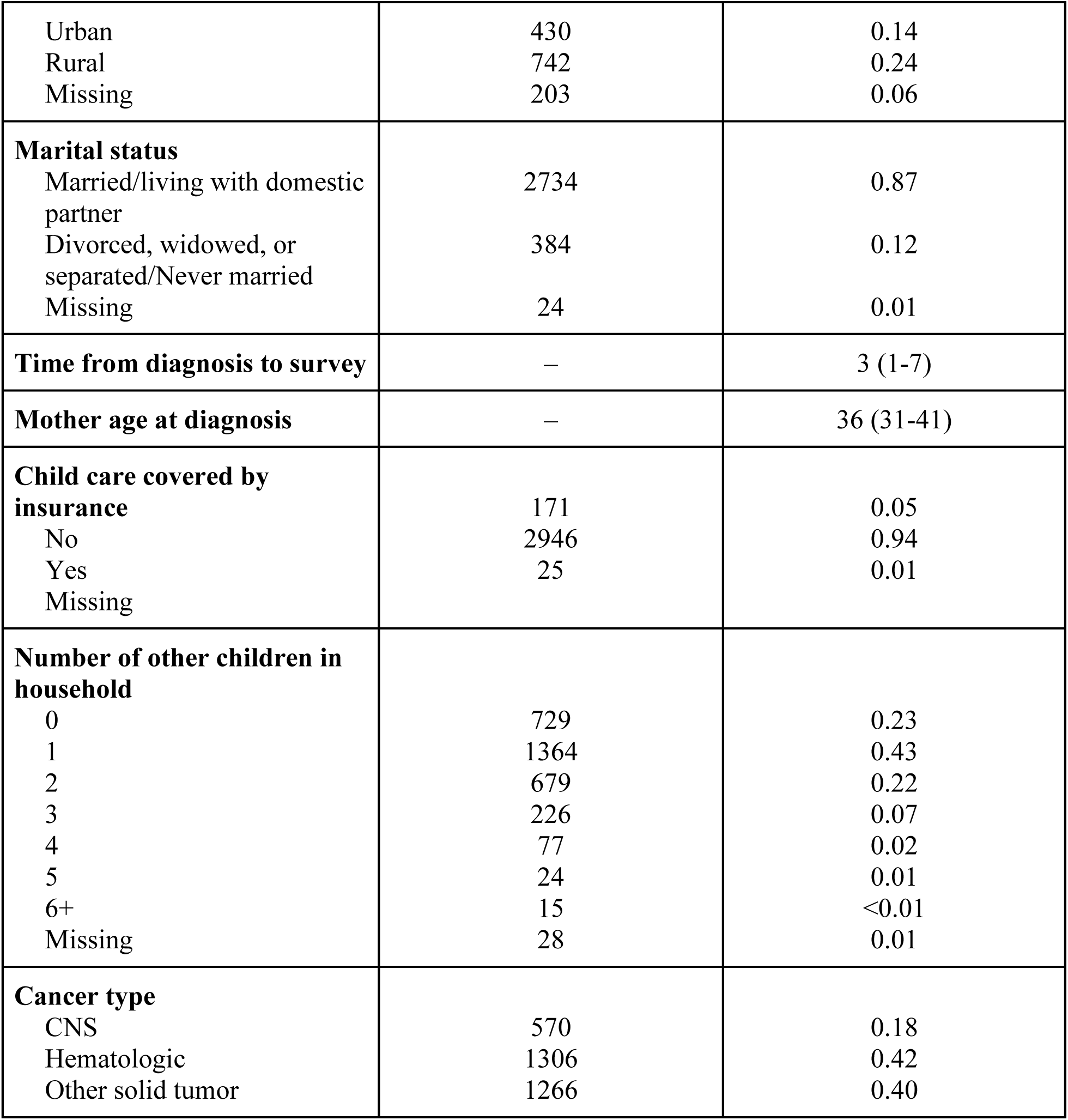
Demographic characteristics of survey respondents.

| **Table 1.** Demographic characteristics of survey respondents |  |  |
| --- | --- | --- |
|  | Number of respondents (n = 3142) | Proportion of study population or median (IQR) |
| **Respondent gender** |  |  |
| Female | 2948 | 0.94 |
| Male | 193 | 0.061 |
| Missing | 1 | <0.01 |
| **Respondent race/ethnicity** |  |  |
| Non-Hispanic white | 2809 | 0.89 |
| Other | 333 | 0.11 |
| **Respondent biological parent** | 3088 | 0.98 |
| **Household education** |  |  |
| Didn't complete high school | 25 | <0.01 |
| Completed high school | 738 | 0.23 |
| Completed college | 905 | 0.29 |
| Completed postgraduate degree | 352 | 0.11 |
| Missing | 1122 | 0.36 |
| **Child gender** |  |  |
| Female | 1421 | 0.45 |
| Male | 1720 | 0.55 |
| Missing | 1 | <0.01 |
| **Child age at diagnosis** | — | 4 (2-9) |
| **Only child vs siblings** |  |  |
| Only child | 729 | 0.23 |
| Has siblings | 2385 | 0.76 |
| Missing | 28 | 0.01 |
| **Household income** |  |  |
| <\$50,000 | 940 | 0.30 |
| \$50,000-\$99,999 | 1275 | 0.41 |
| \$100,000+ | 765 | 0.24 |
| Prefer not to say | 140 | 0.04 |
| Missing | 22 | 0.01 |
| **Locale** |  |  |
| Suburban | 1767 | 0.56 |
| Urban | 430 | 0.14 |
| Rural | 742 | 0.24 |
| Missing | 203 | 0.06 |
| **Marital status** |  |  |
| Married/living with domestic partner | 2734 | 0.87 |
| Divorced, widowed, or separated/Never married | 384 | 0.12 |
| Missing | 24 | 0.01 |
| **Time from diagnosis to survey** | – | 3 (1-7) |
| **Mother age at diagnosis** | – | 36 (31-41) |
| **Child care covered by insurance** |  |  |
| No | 171 | 0.05 |
| Yes | 2946 | 0.94 |
| Missing | 25 | 0.01 |
| **Number of other children in household** |  |  |
| 0 | 729 | 0.23 |
| 1 | 1364 | 0.43 |
| 2 | 679 | 0.22 |
| 3 | 226 | 0.07 |
| 4 | 77 | 0.02 |
| 5 | 24 | 0.01 |
| 6+ | 15 | <0.01 |
| Missing | 28 | 0.01 |
| **Cancer type** |  |  |
| CNS | 570 | 0.18 |
| Hematologic | 1306 | 0.42 |
| Other solid tumor | 1266 | 0.40 |

### Endorsed Emotions in Total Sample

Fear was the most commonly endorsed emotion, with 85% of respondents selecting fear as an emotion that reflected their feelings at the time of their child’s cancer diagnosis. Other emotions selected by a majority of respondents included: worry (69%), sadness (67%), and powerlessness (55%) (**Table 2**).

**Table 2.** Emotions selected in the KMeans clusters.

|  | Proportion of<br>Total Cohort<br>(n = 3142) | Proportion<br>of Cluster 1<br>(n = 1370) | Proportion<br>of Cluster 2<br>(n = 642) | Proportion<br>of Cluster 3<br>(n = 1130) | Chi-square p-value |
| --- | --- | --- | --- | --- | --- |
| **Fear** | 0.85 | 0.89 | 0.87 | 0.80 | 1.4e-09 |
| **Worry** | 0.69 | 0.88 | 0.07 | 0.81 | <2.2e-16 |
| **Sadness** | 0.67 | 0.86 | 0.36 | 0.62 | <2.2e-16 |
| **Powerlessness** | 0.55 | 0.77 | 0.10 | 0.54 | <2.2e-16 |
| **Numbness** | 0.47 | 0.64 | 0.07 | 0.49 | <2.2e-16 |
| **Grief** | 0.44 | 0.64 | 0.23 | 0.32 | <2.2e-16 |
| **Hope** | 0.42 | 0.19 | 0.33 | 0.75 | <2.2e-16 |
| **Anger** | 0.41 | 0.65 | 0.10 | 0.29 | <2.2e-16 |
| **Frustration** | 0.33 | 0.49 | 0.12 | 0.26 | <2.2e-16 |
| **Guilt** | 0.32 | 0.48 | 0.12 | 0.25 | <2.2e-16 |
| **Despair** | 0.29 | 0.49 | 0.17 | 0.12 | <2.2e-16 |
| **Insecurity** | 0.27 | 0.22 | 0.56 | 0.16 | <2.2e-16 |
| **Doubt** | 0.25 | 0.30 | 0.36 | 0.12 | <2.2e-16 |
| **Impatience** | 0.24 | 0.15 | 0.63 | 0.13 | <2.2e-16 |
| **Strength** | 0.24 | 0.05 | 0.22 | 0.49 | <2.2e-16 |
| **Belief** | 0.23 | 0.05 | 0.46 | 0.30 | <2.2e-16 |
| **Loneliness** | 0.22 | 0.33 | 0.03 | 0.18 | <2.2e-16 |
| **Pessimism** | 0.21 | 0.07 | 0.82 | 0.03 | <2.2e-16 |
| **Discouragement** | 0.20 | 0.16 | 0.54 | 0.05 | <2.2e-16 |
| **Appreciation** | 0.17 | 0.07 | 0.20 | 0.27 | <2.2e-16 |
| **Relief** | 0.17 | 0.01 | 0.72 | 0.06 | <2.2e-16 |
| **Blame** | 0.15 | 0.23 | 0.10 | 0.08 | <2.2e-16 |
| **Calm** | 0.15 | 0.01 | 0.52 | 0.12 | <2.2e-16 |
| **Irritation** | 0.15 | 0.12 | 0.36 | 0.05 | <2.2e-16 |
| **Optimism** | 0.15 | 0.05 | 0.01 | 0.35 | <2.2e-16 |
| **Resentment** | 0.15 | 0.20 | 0.19 | 0.05 | <2.2e-16 |
| **Disappointment** | 0.12 | 0.17 | 0.05 | 0.10 | 9.6e-16 |
| **Enthusiasm** | 0.04 | <0.01 | 0.17 | 0.02 | <2.2e-16 |
| **Empowerment** | 0.03 | 0.01 | 0.03 | 0.05 | 8.5e-08 |
| **Passion** | 0.03 | 0.01 | 0.02 | 0.06 | 5.8e-13 |
| **Contentment** | 0.02 | <0.01 | 0.07 | 0.01 | <2.2e-16 |
| **Joy** | 0.01 | <0.01 | 0.01 | 0.03 | 5.1e-10 |

### Dimensionality Reduction

In principal component analysis (PCA) of selected emotions, the first three PCs captured 26.4% of variation. There was a substantial decline in the proportion of variation explained by subsequent PCs, with PCs 4-10 together accounting for only an additional 23% of variation in the data (**Figure 1**). We conducted t-distributed stochastic neighbor embedding (t-SNE) analysis on the selected/unselected emotions of each respondent using a perplexity ranging from 10-50 and – based on PCA results – visualized the data in three dimensions (**Figure 2**). The 3-D t-SNE data generated with perplexity set to 50 were fed into a K-means clustering analysis to identify clusters and to assign respondents to membership within clusters visualized by the t-SNE analysis. Based on the elbow plot method, the optimal K-means value was determined to be three clusters (**Figures 3 and 4**). The proportion of the 3142 respondents categorized into each cluster ranged from 20% (Cluster 2) to 44% (Cluster 1), suggesting reasonable representation of each cluster within the dataset and absence of any small clusters that could have undue influence on downstream analyses.

**Figure 1.**
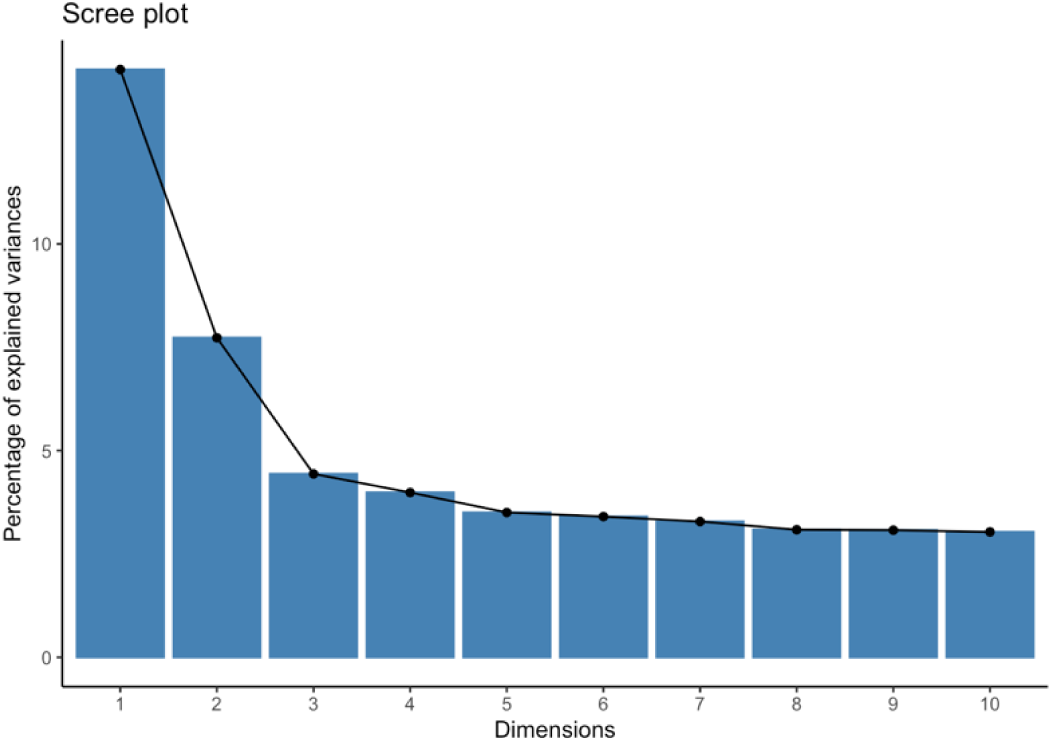
Scree plot of Principal Component Analysis.

**Figure 2.**
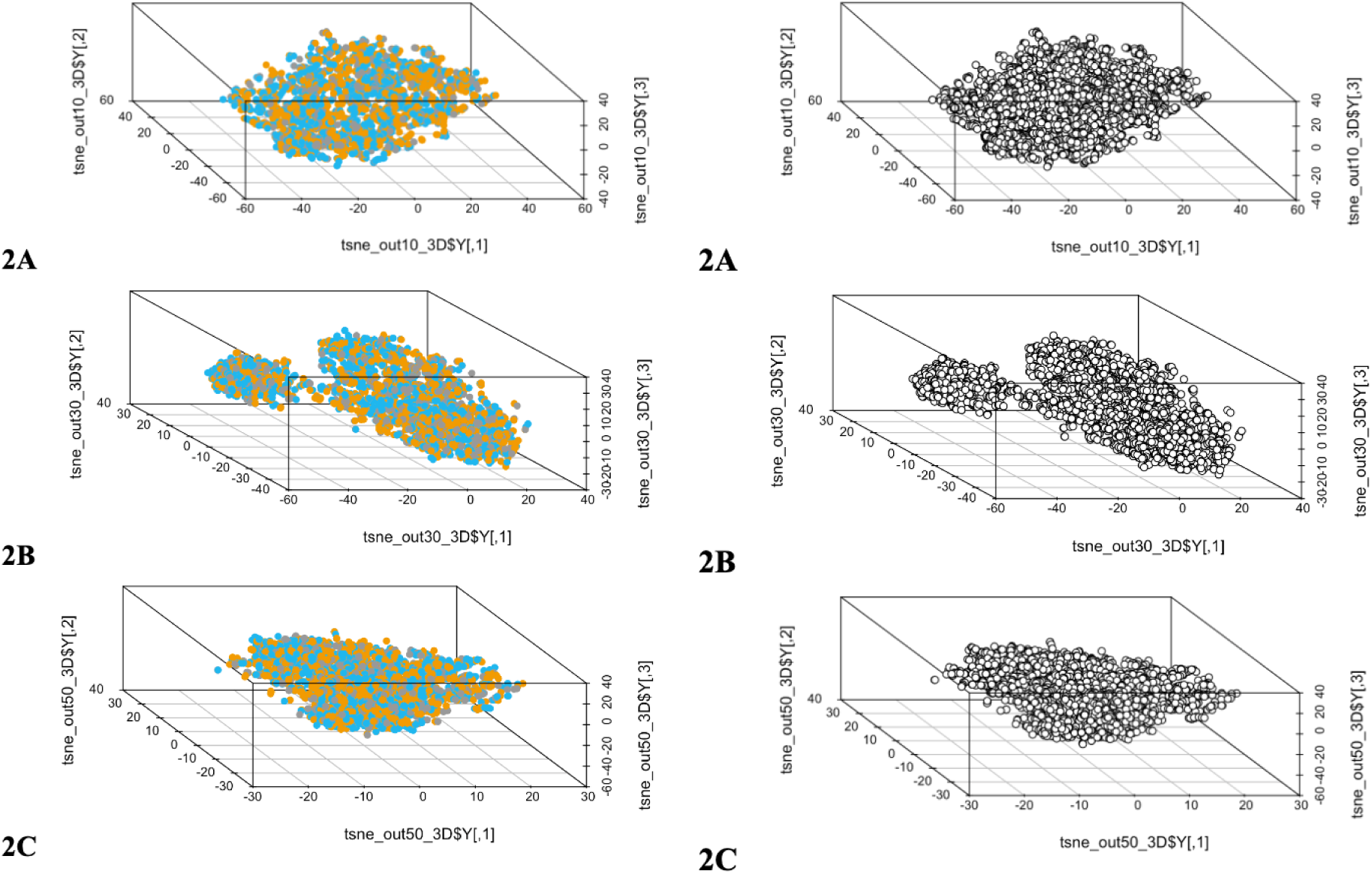
t-SNE Analysis in Three Dimensions. 2A is perplexity = 10, 2B is perplexity = 30, 2C is perplexity = 50.

**Figure 3.**
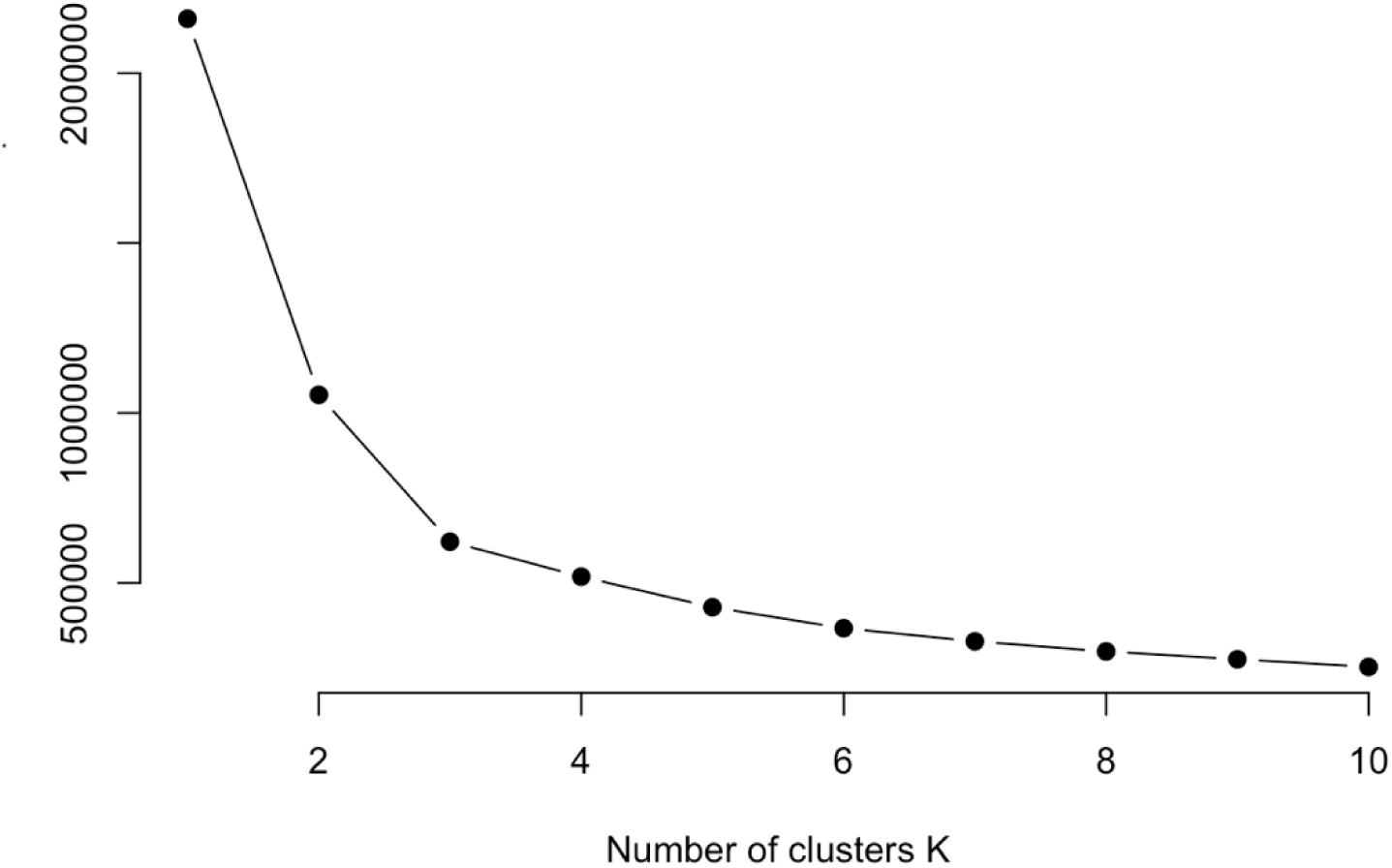
Elbow plot of KMeans clusters. K=1 through K=10, with the sum of squared distances of each point from its centroid plotted on Y axis and K on the X axis.

**Figure 4.**
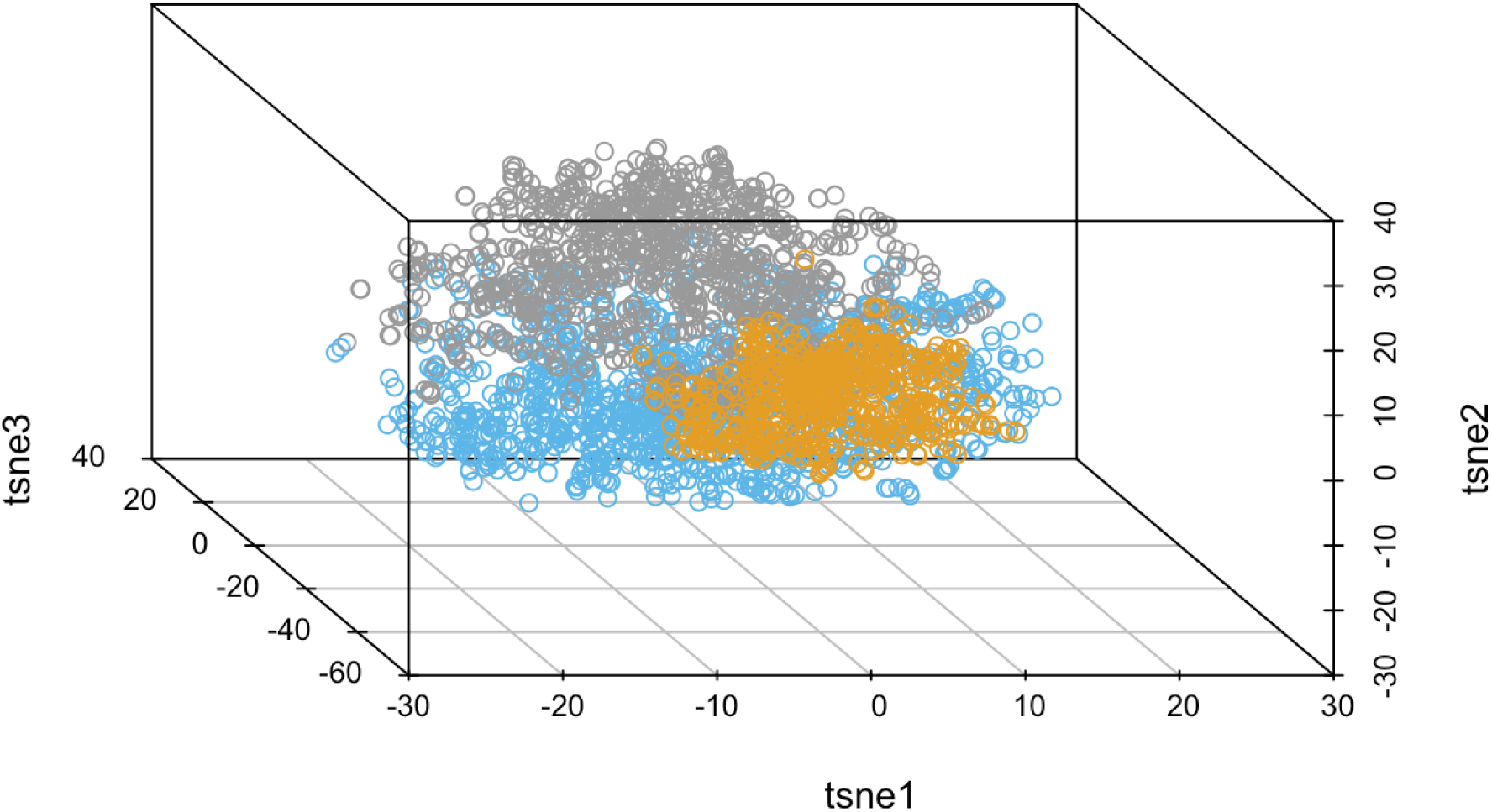
KMeans clustering with t-SNE in 3 dimensions. Blue circles indicate cluster 1, orange circles indicate cluster 2, and grey circles indicate cluster 3.

### Association Between K-means Clusters and Individual Emotions

With all respondents classified into one of three clusters based on the emotions experienced at the time of their child’s cancer diagnosis, we explored the association of each cluster with the 32 individual emotions. As expected based on our neighbor-embedding approach, we observed substantial differences across clusters in the proportion of respondents endorsing various emotions (**Table 2**), which we have visualized in the form of a heatmap (**Figure 5**). Fear – the most endorsed emotion overall – was selected by ≥80% of respondents across all clusters. Worry – the second most endorsed emotion across all respondents – showed substantial heterogeneity across clusters. Only 7% of respondents in Cluster 2 endorsed it, compared to >80% of Clusters 1 and 3. Respondents in Clusters 1 and 3 were also more likely than those in Cluster 2 to endorse worry, sadness, powerlessness, and numbness.

**Figure 5.**
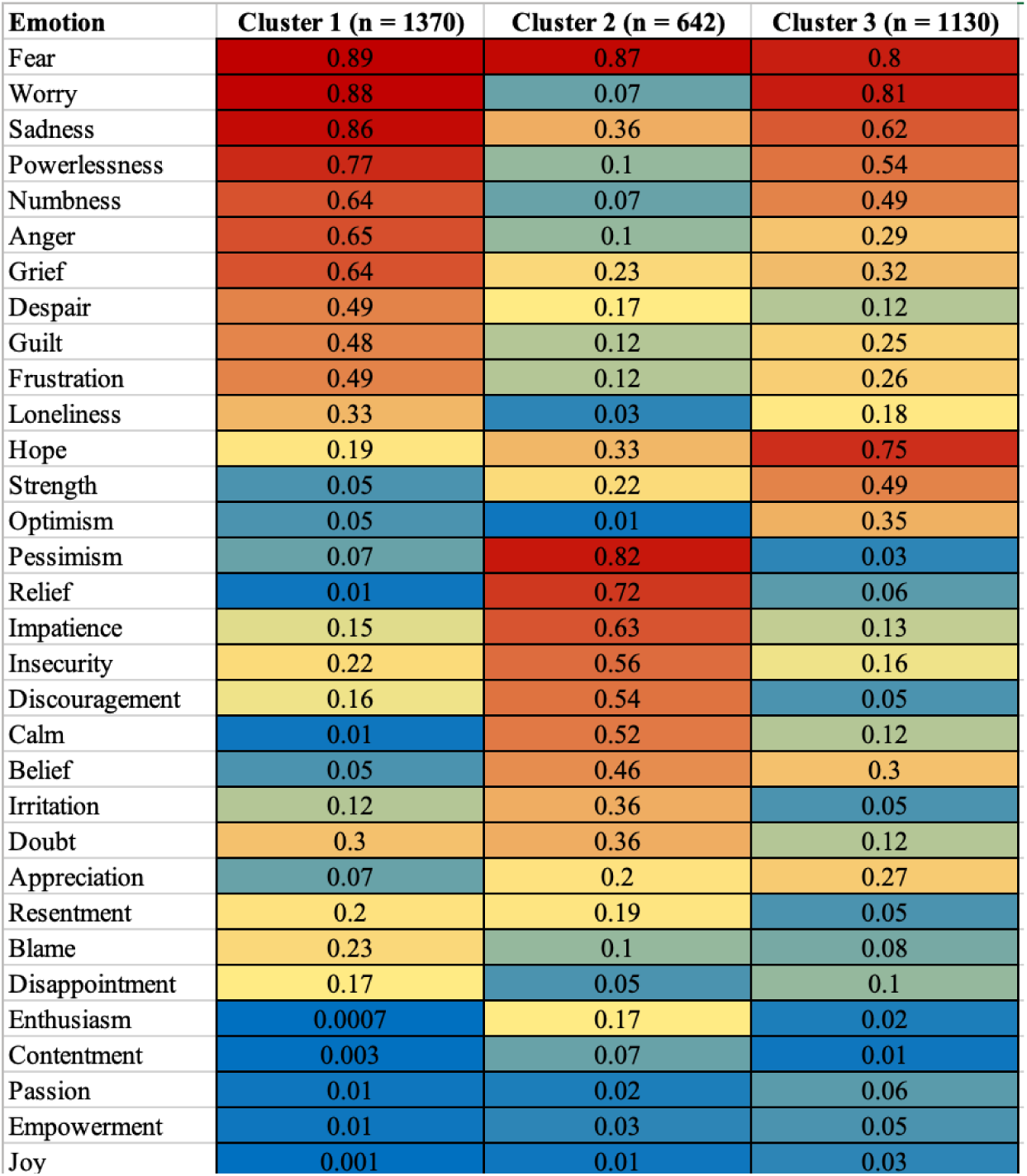
Heat-map of emotions selected in each KMeans cluster. Numbers indicate the proportion of respondents within the cluster that selected the given emotion.

In Cluster 1, anger (65%), grief (64%), despair (49%), frustration (49%), guilt (48%), and loneliness (33%) were endorsed at 3-fold higher levels than in Cluster 2, and at 2-fold higher levels than in Cluster 3. Cluster 2 was uniquely high in its endorsement of pessimism (82%), relief (72%), impatience (63%), insecurity (56%), discouragement (54%), and calm (52%), relative to both Cluster 1 and Cluster 3 (in which these emotions were endorsed at frequencies ranging from 1% to 22%). Cluster 3 was likelier to endorse a number of positive emotions, including hope (75%, compared to 19% in Cluster 1 and 33% in Cluster 2), strength (49% compared to 5% in Cluster 1 and 22% in Cluster 2), and optimism (35% compared to 5% in Cluster 1 and 1% in Cluster 2). This was despite endorsing worry and sadness at frequencies comparable to or higher than that in the other clusters. A subset of emotions was not highly endorsed (≤20%) in any single cluster, including empowerment, passion, contentment, joy, enthusiasm, resentment, and disappointment, although their endorsement frequencies still differed significantly across the clusters (**Table 2**).

### Association Between Emotion Clusters and Demographic/Psychosocial Variables

We explored the association of cluster membership with demographic, clinical and family-structure variables from the diagnosis survey (**Table 3**). Whether or not the respondent was the biological parent of the child differed significantly across clusters, at 99% biological parent in clusters 1 and 2 but 97% in cluster 3 (P=0.013). In terms of household education, Cluster 2 also had higher percentages of respondents that completed college or completed a post-graduate degree (47% and 21%, respectively) (P=4.8×10^-4^). Median child age at diagnosis was 3 in Cluster 2 and 5 in Clusters 1 and 3 (P=2.0×10^-6^). While all the clusters had similar percentages of respondents with a household income of $50-99,000 (around 42-43%), Cluster 2 had a lower percentage of respondents with a household income of <$50,000 a year (25% vs 33% in Clusters 1 and 3), and a higher percentage of respondents with a household income of >$100,000 a year (32% vs 25% and 23% in clusters 1 and 3 respectively) (P=4.0×10^-5^). Overall, 42% of respondents’ children were diagnosed with a hematologic cancer, and within clusters, this proportion was 42% for Cluster 1, 38% for Cluster 2, and 43% for Cluster 3 (P=0.059). 18% of total respondents’ children were diagnosed with a CNS malignancy, which within clusters came to 21% in Cluster 1, 16% in Cluster 2, and 16% in Cluster 3 (P=1.1×10^-3^). Finally, while overall 40% of respondents’ children were diagnosed with another solid tumor, within clusters, this proportion was 37% for Cluster 1, 47% for Cluster 2, and 41% for Cluster 3 (P=1.1×10^-4^). Cluster membership did not differ by respondent sex, respondent race/ethnicity, insurance status, marital status, or sibling status.

**Table 3.**
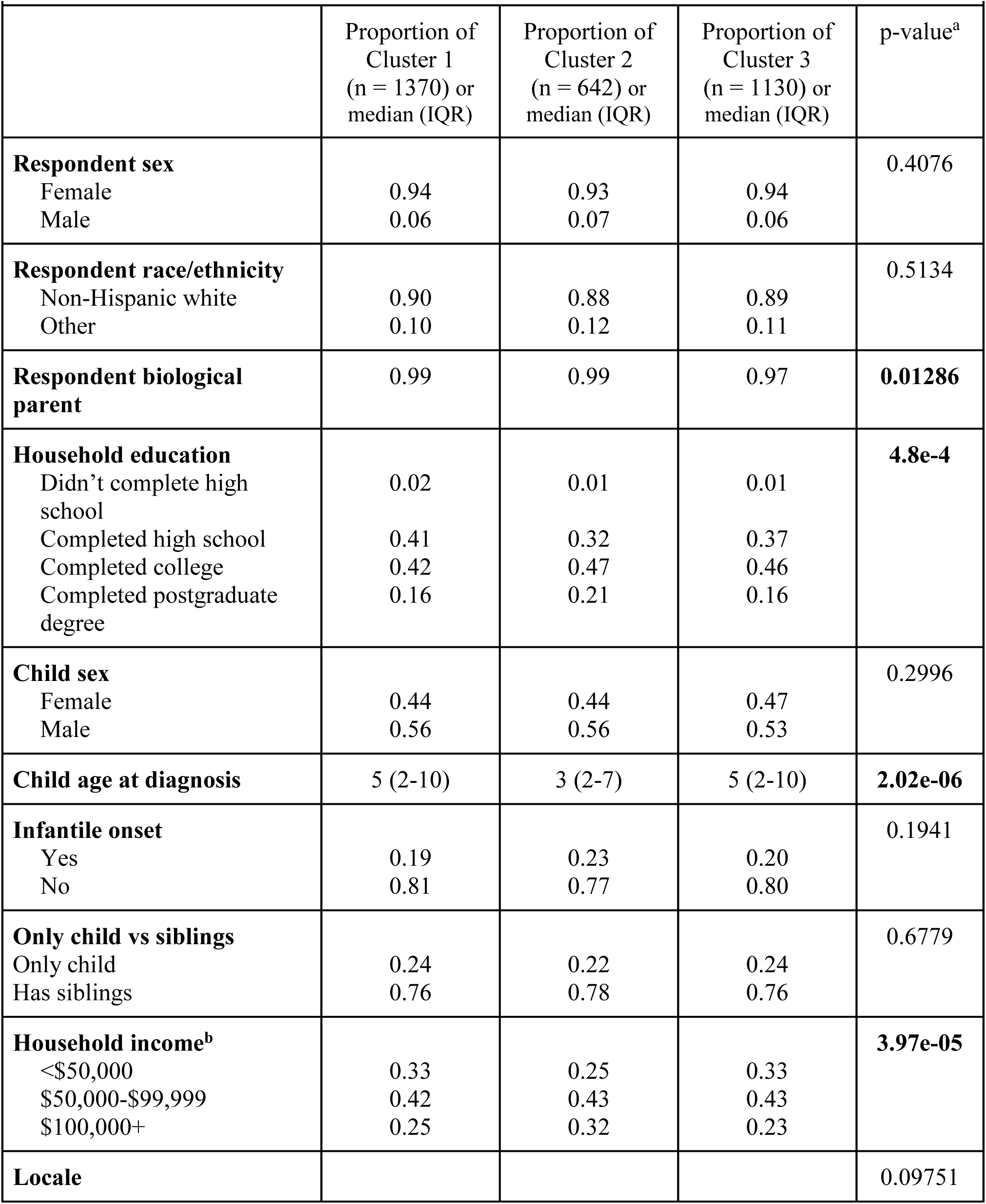
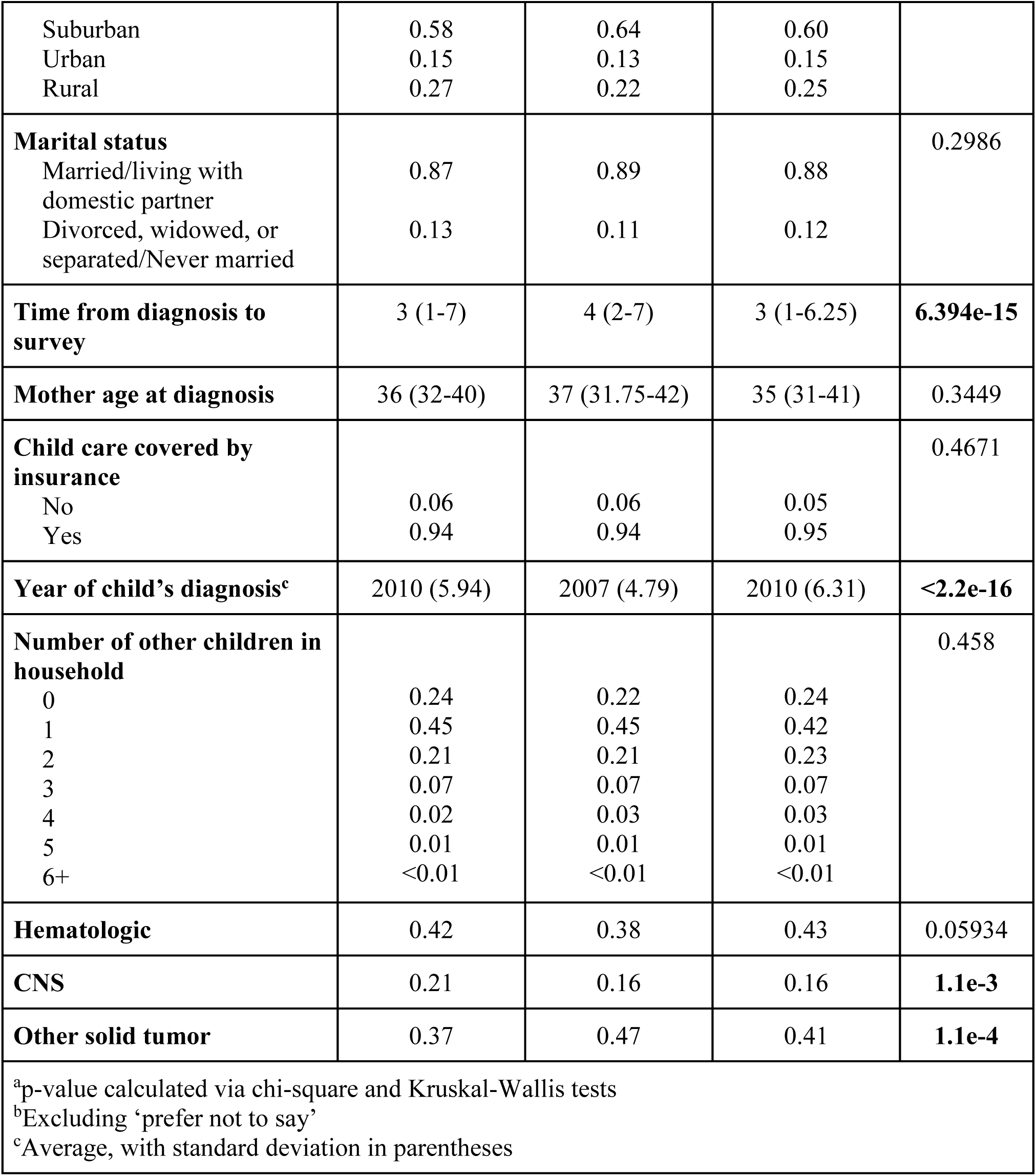
Associations between KMeans clusters and demographic/psychosocial variables.

|  |  |  |  |  |
| --- | --- | --- | --- | --- |
|  | Proportion of<br>Cluster 1<br>(n = 1370) or<br>median (IQR) | Proportion of<br>Cluster 2<br>(n = 642) or<br>median (IQR) | Proportion of<br>Cluster 3<br>(n = 1130) or<br>median (IQR) | p-value<sup>a</sup> |
| **Respondent sex** |  |  |  | 0.4076 |
| Female | 0.94 | 0.93 | 0.94 |  |
| Male | 0.06 | 0.07 | 0.06 |  |
| **Respondent race/ethnicity** |  |  |  | 0.5134 |
| Non-Hispanic white | 0.90 | 0.88 | 0.89 |  |
| Other | 0.10 | 0.12 | 0.11 |  |
| **Respondent biological parent** | 0.99 | 0.99 | 0.97 | **0.01286** |
| **Household education** |  |  |  | **4.8e-4** |
| Didn't complete high school | 0.02 | 0.01 | 0.01 |  |
| Completed high school | 0.41 | 0.32 | 0.37 |  |
| Completed college | 0.42 | 0.47 | 0.46 |  |
| Completed postgraduate degree | 0.16 | 0.21 | 0.16 |  |
| **Child sex** |  |  |  | 0.2996 |
| Female | 0.44 | 0.44 | 0.47 |  |
| Male | 0.56 | 0.56 | 0.53 |  |
| **Child age at diagnosis** | 5 (2-10) | 3 (2-7) | 5 (2-10) | **2.02e-06** |
| **Infantile onset** |  |  |  | 0.1941 |
| Yes | 0.19 | 0.23 | 0.20 |  |
| No | 0.81 | 0.77 | 0.80 |  |
| **Only child vs siblings** |  |  |  | 0.6779 |
| Only child | 0.24 | 0.22 | 0.24 |  |
| Has siblings | 0.76 | 0.78 | 0.76 |  |
| **Household income<sup>b</sup>** |  |  |  | **3.97e-05** |
| <\$50,000 | 0.33 | 0.25 | 0.33 | |
| \$50,000-\$99,999 | 0.42 | 0.43 | 0.43 | |
| \$100,000+ | 0.25 | 0.32 | 0.23 | |
| **Locale** |  |  |  | 0.09751 |
| Suburban | 0.58 | 0.64 | 0.60 |  |
| Urban | 0.15 | 0.13 | 0.15 |  |
| Rural | 0.27 | 0.22 | 0.25 |  |
| **Marital status** |  |  |  | 0.2986 |
| Married/living with domestic partner | 0.87 | 0.89 | 0.88 |  |
| Divorced, widowed, or separated/Never married | 0.13 | 0.11 | 0.12 |  |
| **Time from diagnosis to survey** | 3 (1-7) | 4 (2-7) | 3 (1-6.25) | **6.394e-15** |
| **Mother age at diagnosis** | 36 (32-40) | 37 (31.75-42) | 35 (31-41) | 0.3449 |
| **Child care covered by insurance** |  |  |  | 0.4671 |
| No | 0.06 | 0.06 | 0.05 |  |
| Yes | 0.94 | 0.94 | 0.95 |  |
| **Year of child's diagnosis<sup>c</sup>** | 2010 (5.94) | 2007 (4.79) | 2010 (6.31) | **<2.2e-16** |
| **Number of other children in household** |  |  |  | 0.458 |
| 0 | 0.24 | 0.22 | 0.24 |  |
| 1 | 0.45 | 0.45 | 0.42 |  |
| 2 | 0.21 | 0.21 | 0.23 |  |
| 3 | 0.07 | 0.07 | 0.07 |  |
| 4 | 0.02 | 0.03 | 0.03 |  |
| 5 | 0.01 | 0.01 | 0.01 |  |
| 6+ | <0.01 | <0.01 | <0.01 |  |
| **Hematologic** | 0.42 | 0.38 | 0.43 | 0.05934 |
| **CNS** | 0.21 | 0.16 | 0.16 | **1.1e-3** |
| **Other solid tumor** | 0.37 | 0.47 | 0.41 | **1.1e-4** |
| <sup>a</sup>p-value calculated via chi-square and Kruskal-Wallis tests<br><sup>b</sup>Excluding 'prefer not to say'<br><sup>c</sup>Average, with standard deviation in parentheses |  |  |  |  |

The median time from diagnosis to survey completion was longest in Cluster 2 (4, P=6.4×10^-15^), and the average year of the child’s diagnosis was earliest in Cluster 2 (2007, P=<2.2×10^-16^).

## DISCUSSION

This study sought to explore variation in the emotional state of parents/caregivers at the time of a child’s cancer diagnosis and evaluate how the emotional response was associated with certain sociodemographic, clinical, and familial factors. The endorsement of any one emotion was strongly correlated (both positively and negatively) with the endorsement of the 31 other remaining emotions. This complex inter-dependence led us to perform dimensionality reduction, identifying three clusters of emotional responses that were well-represented across the data set and captured 44%, 20%, and 36% of respondents, respectively. Each of the 32 emotions appeared to contribute to the clustering analyses, as each was endorsed at significantly different frequencies across clusters. However, the emotions were not neatly partitioned across clusters in a manner that facilitates discrete assignment of a descriptive label to each cluster.

Despite substantive overlap in emotions endorsed within and across clusters, we identified several “hallmark” emotions endorsed by >50% of respondents within a single cluster and by <35% of respondents in the other two clusters, including: “anger and grief” (Cluster 1); “pessimism, relief, impatience, insecurity, discouragement, and calm” (Cluster 2); and “hope” (Cluster 3). Also of note, Cluster 2 had <35% endorsement of “worry and powerlessness” while Clusters 1 and 3 endorsed these emotions at frequencies >50%, suggesting that the absence of worry and powerlessness were additional hallmarks of membership in Cluster 2. As an aide to discussion, the remainder of the paper will reference these hallmark emotions when referring to specific clusters. However, we emphasize that the hallmark emotions are in no way unique to any one cluster, nor is there strong evidence that they are necessarily the prime drivers of cluster membership.

Cluster 1 (“anger and grief”) had the lowest endorsement of both optimism and pessimism which, despite being antonyms, are future-focused emotions focused. Similarly, this cluster was most likely to endorse both guilt and blame, both of which are emotions focused on past events. Although enriched for several traditionally negative emotions compared to other clusters, Cluster 2 (“pessimism, relief, impatience, insecurity, discouragement, and calm”) had lower endorsement of several other negative emotions, compared to Cluster 1, including despair, frustration, powerlessness, and worry. The high levels of pessimism and impatience, combined with low levels of despair and powerlessness, may reflect more active emotional states in Cluster 2, compared with the more dormant emotional states in Cluster 1.^17^ Cluster 3 (“hope”) endorsed a number of traditionally positive emotions at higher frequencies than other clusters, most notably hope, but also appreciation, strength, and optimism. Despite this, Cluster 3 appears cognizant of the seriousness of their child’s diagnosis, with a majority of respondents also endorsing fear, worry, and sadness.

Importantly, cluster membership was associated with differences in a number of parental factors, including educational attainment, family income, and status as the biological parent. Equally interesting, some parental factors were unassociated with cluster membership, the most notable of which was respondent sex. While other analyses have identified notable differences in the psychosocial wellbeing of mothers versus fathers of children with cancer, such studies have focused on well-being during the course of the child’s treatment and convalescence,^18,19^ or following a child’s death from cancer,^20^ and are likely influenced by differences in caregiving responsibilities and return-to-work behaviors. Our study is unique is comparing emotions experienced at the time of diagnosis, where robust sex-differences were not observed.

Cluster membership was also associated with child-specific factors, including age at diagnosis and cancer type. Children of parents in Cluster 2 (“pessimism, relief, impatience, insecurity, discouragement, and calm”) were significantly younger than children of parents in other clusters, which may contribute to experiencing both fewer positive emotions than Cluster 3 and experiencing more “active” emotions than Cluster 1. CNS tumors were most commonly diagnosed in Cluster 1 families (“anger and grief”), other solids tumors in Cluster 2 families (“pessimism, relief, impatience, insecurity, discouragement, and calm”), and hematologic malignancies in Cluster 3 families (“hope”). Differences in both overall prognosis and in the invasiveness of therapeutic modalities across cancer types seems likely to underlie some of these differences, with parents of children with CNS tumors experiencing negative emotions that look more to the past than the future and parents of children with hematologic malignancies likelier to endorse hope, strength, and optimism.

A cross-sectional study assessing anticipatory grief of caregivers of children with cancer in Jordan found that parents of newly diagnosed children reported high levels of personal sacrifice burden, “terrific sadness,” and worry for the future.^3^ Similar results were observed in our data, as well as in a cross-sectional study conducted in China,^5^ reflecting cross-cultural similarities in caregiver emotional responses. However, our results also differ from prior literature in important ways. While worry and sadness were highly endorsed overall (69% and 67%, respectively), they had low endorsement in Cluster 2 which suggests greater heterogeneity in emotional responses than traditionally appreciated. The low endorsement of grief in our sample (44%) also suggests that pathways toward experiencing anticipatory grief among parents of children with cancer may take time to manifest and may be influenced by both caregiver and child-level factors. Other international studies conducted in Brazil, Chile, and New Zealand analyzing the impact of childhood cancer on parent mental health observed relationships between higher stress/anxiety and the age of the parent, fears of a negative prognosis, difficulties with familial relationships, and financial hardship.^6,7,8,9^ We observed that emotional responses were associated with both education and household income, supporting roles for financial and educational differences in influencing how parents understand and respond to a child’s diagnosis.

Relative to other studies investigating caregiver emotions, our study has a very large sample, which partially motivated our approach to assessing parental emotional responses. By asking respondents to select representative emotions from a large list randomized to each participant at survey delivery, we limit *a priori* assumptions about the breadth and diversity of the emotions that may be experienced by parents of children with cancer. While validated scales exist for depression, anxiety, and grief, the approach taken in this exploratory analysis permits the incorporation of additional potentially relevant emotions such as loneliness, hope, and strength. Further, the large number of emotions included in the survey were made analytically manageable through rigorous dimensionality reduction techniques.

Our study also has several important limitations that merit consideration. First, survey participants represent a self-selected population of caregivers who independently navigated to the ALSF MCC survey portal. While a random sampling of childhood cancer caregivers would be optimal, this is generally infeasible in practice and most studies to-date involve subjects recruited at a single institution – typically a tertiary referral center. Our study was cross-sectional in nature, and relies on retrospective recall of emotions experienced at time of diagnosis. Future work in the peri-diagnostic setting could address this issue, but requires caution given the potentially fragile emotional state of parents at that time and their focus on more pressing matters than psychosocial surveys. Survey respondents were relatively homogenous in terms of both sex and race/ethnicity. Study results may not translate to other populations, although prior research has found high cross-cultural concordance in parental emotional responses which may minimize this issue.

Parental well-being is critical to child care, particularly when a family faces such devastating situations like a cancer diagnosis. The results of this study indicate substantial heterogeneity in emotional responses following a child’s cancer diagnosis, with differences related to both caregiver-level and child-level factors. Recognizing these differences will be important in developing better approaches to support caregivers and their families at the time of diagnosis, with an understanding that there is no “right” or “wrong” way to feel when faced with a child’s cancer diagnosis. Initiating responsive and effective programs at the time of diagnosis will help to provide targeted and adaptive support for parents throughout their family’s cancer journey.

## Data Availability

De-identified, individual-level data are available from the authors upon reasonable request.

## Abbreviations

ALSF: Alex’s Lemonade Stand Foundation
CNS: central nervous system
MCC: My Childhood Cancer
PCA: Principal component analysis
t-SNE: t-stochastic neighbor embedding

## REFERENCES

1. US Childhood Cancer Statistics. (n.d.). ACCO. Retrieved November 13, 2021, from https://www.acco.org/us-childhood-cancer-statistics/

2. Childhood Cancer Facts. (n.d.). Retrieved November 13, 2021, from https://www.stjude.org/treatment/pediatric-oncology/childhood-cancer-facts.html

3. Al-Gamal, E., & Long, T. (2010). Anticipatory Grieving Among Parents Living with a Child with Cancer. Journal of Advanced Nursing, 66(9), 1980–1990. https://doi.org/10.1111/j.1365-2648.2010.05381.x

4. Cardinali P, Migliorini L, & Rania N. (2019). The Caregiving Experiences of Fathers and Mothers of Children with Rare Diseases in Italy: Challenges and Social Support Perceptions. Frontiers in Psychology, 10(1780). https://doi.org/10.3389/fpsyg.2019.01780

5. Yu, W., Lu, Q., Lu, Y., Yang, H., Zhang, L., Guo, R., & Hou, X. (2021). Anticipatory Grief Among Chinese Family Caregivers of Patients with Advanced Cancer: A Cross-Sectional Study. Asia-Pacific Journal of Oncology Nursing, 8(4), 369–376. https://doi.org/10.4103/apjon.apjon-214

6. Alves, D. F. dos S., Guirardello, E. de B., & Kurashima, A. Y. (2013). Stress Related to Care: The Impact of Childhood Cancer on the Lives of Parents. Revista Latino-Americana de Enfermagem, 21, 356–362. https://doi.org/10.1590/S0104-11692013000100010

7. Borrescio-Higa, F., & Valdés, N. (2022). The Psychosocial Burden of Families with Childhood Blood Cancer. International Journal of Environmental Research and Public Health, 19(1), 599. https://doi.org/10.3390/ijerph19010599

8. Dockerty, J. D., Williams, S. M., McGee, R., & Skegg, D. C. G. (2000). Impact of Childhood Cancer on the Mental Health of Parents. Medical and Pediatric Oncology, 35(5), 475–483. https://doi.org/10.1002/1096-911X(20001101)35:5<475::AID-MPO6>3.0.CO;2-U

9. Rosenberg, A. R., Dussel, V., Kang, T., Geyer, J. R., Gerhardt, C. A., Feudtner, C., & Wolfe, J. (2013). Psychological Distress in Parents of Children with Advanced Cancer. JAMA Pediatrics, 167(6), 537–543. https://doi.org/10.1001/jamapediatrics.2013.628

10. Bogetz, J. F., Trowbridge, A., Kingsley, J., Taylor, M., Rosenberg, A. R., & Barton, K. S. (2020). “It’s My Job to Love Him”: Parenting Adolescents and Young Adults With Advanced Cancer. Pediatrics, 146(6). https://doi.org/10.1542/peds.2020-006353

11. Bogetz, J., Trowbridge, A., Kingsley, J., Taylor, M., Wiener, L., Rosenberg, A. R., & Barton, K. S. (2021). Stuck Moments and Silver-Linings: The Spectrum of Adaptation Among Non-Bereaved and Bereaved Parents of Adolescents and Young Adults With Advanced Cancer. Journal of Pain and Symptom Management, 62(4), 709–719. https://doi.org/10.1016/j.jpainsymman.2021.03.015

12. Wimberly C. E., Towry L., Caudill C., Johnston E. E., & Walsh K. M. (2021). Impacts of COVID-19 on Caregivers of Childhood Cancer Survivors. Pediatric Blood Cancer, 68(4), e28943. https://www.ncbi.nlm.nih.gov/pmc/articles/PMC7995053/

13. Krijthe, J.H. (2015). *Rtsne: T-Distributed Stochastic Neighbor Embedding Using Barnes-Hut Implementation*. R package version 0.15, https://github.com/jkrijthe/Rtsne

14. Ligges, U. & Mächler, M. (2003). Scatterplot3d – an R Package for Visualizing Multivariate Data. Journal of Statistical Software 8(11), 1–20.

15. Venables W.N., & Ripley B.D. (2002). *Modern Applied Statistics with S*, Fourth edition. Springer, New York. ISBN 0-387-95457-0, https://www.stats.ox.ac.uk/pub/MASS4/.

16. R Core Team (2021). R: A language and environment for statistical computing. R Foundation for Statistical Computing, Vienna, Austria. https://www.R-project.org/

17. Stout, R. (2022). Dormant and active emotional states. Synthese 200(161) https://doi.org/10.1007/s11229-022-03652-8

18. Rensen, N., Steur, L. M., Schepers, S. A., Merks, J. H., Moll, A. C., Kaspers, G. J., Grootenhuis, M. A., & van Litsenburg, R. R. (2019). Gender-specific Differences in Parental Health-Related Quality of Life in Childhood Cancer. Pediatric Blood & Cancer, 66(7), e27728. https://doi.org/10.1002/pbc.27728

19. Mogensen, N., Saaranen, E., Olsson, E., Klug Albertsen, B., Lähteenmäki, P. M., Kreicbergs, U., Heyman, M., & Harila-Saari, A. (2022). Quality of Life in Mothers and Fathers of Children Treated for Acute Lymphoblastic Leukaemia in Sweden, Finland and Denmark. British Journal of Haematology, 198(6), 1032–1040. https://doi.org/10.1111/bjh.18350

20. Goodenough, B., Drew, D., Higgins, S., & Trethewie, S. (2004). Bereavement Outcomes for Parents Who Lose a Child to Cancer: Are Place of Death and Sex of Parent Associated with Differences in Psychological Functioning?. Psycho-oncology, 13(11), 779–791. https://doi.org/10.1002/pon.795

